# Using routine laboratory tests to perform early prediction of urine culture results

**DOI:** 10.1101/2025.11.24.25340905

**Authors:** Namrata Harish, Cindy Zhang, Ya-Lin Chen, Brody H Foy

## Abstract

**Background:** Urinary tract infections (UTIs) are among the most common bacterial infections worldwide, typically diagnosed using a urine culture. However, urine cultures can take up to 72hrs to result, delaying and inhibiting treatment decisions. Here we aim to investigate whether machine learning models can enable earlier prediction of urine culture results.

**Methods:** We analyzed 30,369 urine cultures from 10,761 patients in the University of Washington Medicine Network. Random forest models were developed to predict overall culture positivity, as well as presence of each of the 10 most common infectious agents, using patient demographics and routine laboratory tests (blood counts, metabolic panels, etc.). Age- and sex-adjusted univariate associations of lab markers with culture positivity were also analyzed via logistic regression.

**Results:** ML models predicted culture positivity with moderate accuracy (AUC: 0.76), and high precision (80-90%) at clinically relevant recall rates, with higher performance in cases where urinalysis was available (AUC: 0.82). Individual pathogen prediction was somewhat lower (AUCs: 0.64-0.72) though, particularly for less common pathogens. At least one marker from all common test panels showed a significant univariate association with culture results.

**Conclusions:** ML models using routine laboratory tests can predict overall urine culture positivity with clinically useful accuracy, though individual pathogen prediction remains challenging. Given limited urinalysis availability, these models may be best suited for triaging presumptively negative cultures to improve laboratory efficiency rather than directly informing antibiotic selection.

## Introduction

Urinary tract infections (UTIs) are among the most common bacterial infections worldwide, with particularly high prevalence in women and older adults^1–3^. They are typically driven by pathogens such as Escherichia coli and klebsiella pneumoniae, and can lead to wide ranging symptoms from mild discomfort to severe systemic illness^1^. Diagnosis and management of UTIs requires a careful balance between prompt treatment to prevent complications, and judicious antibiotic use to minimize resistance development^4^. This is particularly true for vulnerable populations such as those with recurrent infections or immunosuppressed systems, where delayed or inappropriate treatment can lead to serious consequences.

To enable accurate diagnosis, the gold standard for UTIs is the urine culture – which can typically confirm the type of underlying pathogen, and potentially its antimicrobial susceptibility profile^4,5^. However, standard culture methods require 24-72hrs for bacterial growth^4,5^, creating a challenging gap between patient presentation and therapy initiation. During this period, clinicians must often decide whether to prescribe antibiotics based on the patient’s history and presentation. This in turn leads to substantial overuse of antibiotics, contributing to the growing global challenge of antibiotic resistance^3,6–8^.

As such, there would be significant value in early prediction of urine culture results, to rapidly inform treatment pathways. In this context, machine learning (ML) algorithms offer substantial promise^9,10^. Simple ML algorithms have been utilized widely across medicine to enable rapid prediction of clinical outcomes and/or improve efficiency of the clinical laboratory^11–13^, suggesting similar approaches may provide value for this use case. However, a major challenge is that most urine cultures are ordered in outpatient and/or family medicine contexts, where substantial additional data (laboratory tests, etc.) is not always available for each patient. Given this, here we aim to investigate whether ML models can enable early prediction of urine culture results, using only routinely collected laboratory tests such as the complete blood count and metabolic panel.

## Methods

### Data collection

We considered all urine culture tests ordered as part of standard clinical care in the University of Washington Medical Network between January 1, 2021 and June 19, 2024. Patients who did not have a complete blood count (CBC) test prior to the first culture result were excluded. This led to 10,761 patients, and 30,369 total cultures. For each patient, all quantitative laboratory test results were taken from two weeks prior to the culture order until time of the cultures first result. If a lab test was measured multiple times, the value closest to the culture order time was taken. Culture tests typically generate many results over time, based on how long the potential pathogens take to grow to a detectable level. For all analyses we took the time of the cultures first positive finding, or for the negative cultures, the time of the last negative report, as the primary result time.

Collected laboratory tests included the complete blood count (hemoglobin [HB], hematocrit [HCT], mean corpuscular hemoglobin [MCH], mean corpuscular hemoglobin concentration [MCHC], mean corpuscular volume [MCV], mean platelet volume [MPVF], platelet count [PLT], red cell count [RBC], red cell distribution width [RDW], and white cell count [WBC]), white cell differential (absolute neutrophil [TNEUT], lymphocyte [LYMPH], eosinophil [EOS], monocyte [MONOC], basophil [BASO] immature granulocytes [IMG], and nucleated red cell counts [NRBC]), the metabolic panel (anion gap [IGAP], blood urea nitrogen [BUN], calcium [CA], chloride [CL], carbon dioxide [CO2], creatinine [CRE], estimated glomerular filtration rate [EGFR], glucose [GLU], potassium [K]), live function tests (albumin [ALB], alkaline phosphatase [ALK], aspartate aminotransferase [AST], alanine aminotransferase [ALT], bilirubin [BIL], total protein [TP]), urinalysis (urine WBC [UWBC], RBC [URBC], pH [UPH], specific gravity [USPG], squamous [USQEP] and renal/transitional epithelial cells [URTEPI]), antibiotic susceptibility testing (ciprofloxacin [CIP], nitrofurantoin [FM], gentamicin [GM], ceftazidime [CAZ], meropenem [MER], cefepime [CEFPM], piperacillin/tazobactam [PIPTAZ], ampicillin [AMP], ertapenem [ERT], aztreonam [ATM], cefazolin [CFZ], ampicillin-sulbactam [SAM], cefotetan [CTT], moxifloxacin [MOXFL], levofloxacin [LEV], trimethoprim-sulfamethoxazole [SXT], and ceftriaxone [TRX]), coagulopathic testing (prothrombin time [PROPAT], prothrombin international normalized ratio [PROINR]).

### Model development

We trained models for two related tasks: prediction of overall urine culture positivity (identification of any infectious agent), and prediction of each of the 10 most common infectious agents (Escherichia coli, klebsiella, enterococcus, streptococcus, staphylococcus, lactobacillus, proteus, pseudomonas, Citrobacter, and yeast). For each task we trained models using demographic data, and any available quantitative laboratory test data. We trained separate models for prediction at 0, 6, 12, 24, 48, and 72 hours post culture collection, limited to samples that had not yet had a positive result for that agent or been finalized as negative. Multi-categorical variables (e.g., race, etc.) were one-hot encoded. Results above or below a reporting limit were cast as that limit (e.g., >100 converted to 100), and missing data was numerically encoded as −1. All data was Z-score normalized prior to model training.

Using this data, each model was trained with an 80:10:10 split into training, validation and test sets at the patient level. We considered logistic regressions, random forests, and gradient boosted tree models^14^ (XGBoost). For each model, we performed hyperparameter optimization using a random grid search method, with validation set area under the receiver-operator characteristic curve (AUC) as the primary performance metric. For each trained model, we calculated feature importance using the Gini impurity metric. All presented results will focus on random forest models, the best performing model.

### Statistical analysis

All data analysis was performed using *Python 3.12*, with model training performed using the *scikit-learn* package. Age- and sex-adjusted odds ratios between laboratory tests and culture results were calculated using logistic regression models with the *statsmodels* package. Correlations were calculated using the Spearman method.

## Results

The primary cohort consisted of 10,761 patients and 30,369 cultures. The cohort was 36.9% male, with mean(std) age of 60.8(18.5)y at time of culture order. Overall, 76% of cultures generated at least one positive finding.

### Data availability and timing

The cumulative distribution of culture result times is given in **Fig. 1a**. Most cultures were first resulted between 24- and 48-hrs, suggesting that result prediction would have the largest clinical utility in the first 24hrs post order. The top positive agent was mixed flora, then E. coli, with all other agents having positivity rates below 10% **(Fig. 1b)**. A mild difference in age was seen between positive and negative cultures (61.0 vs 60.2y, p < 0.001, **Fig. 1c)**.

**Figure 1.**
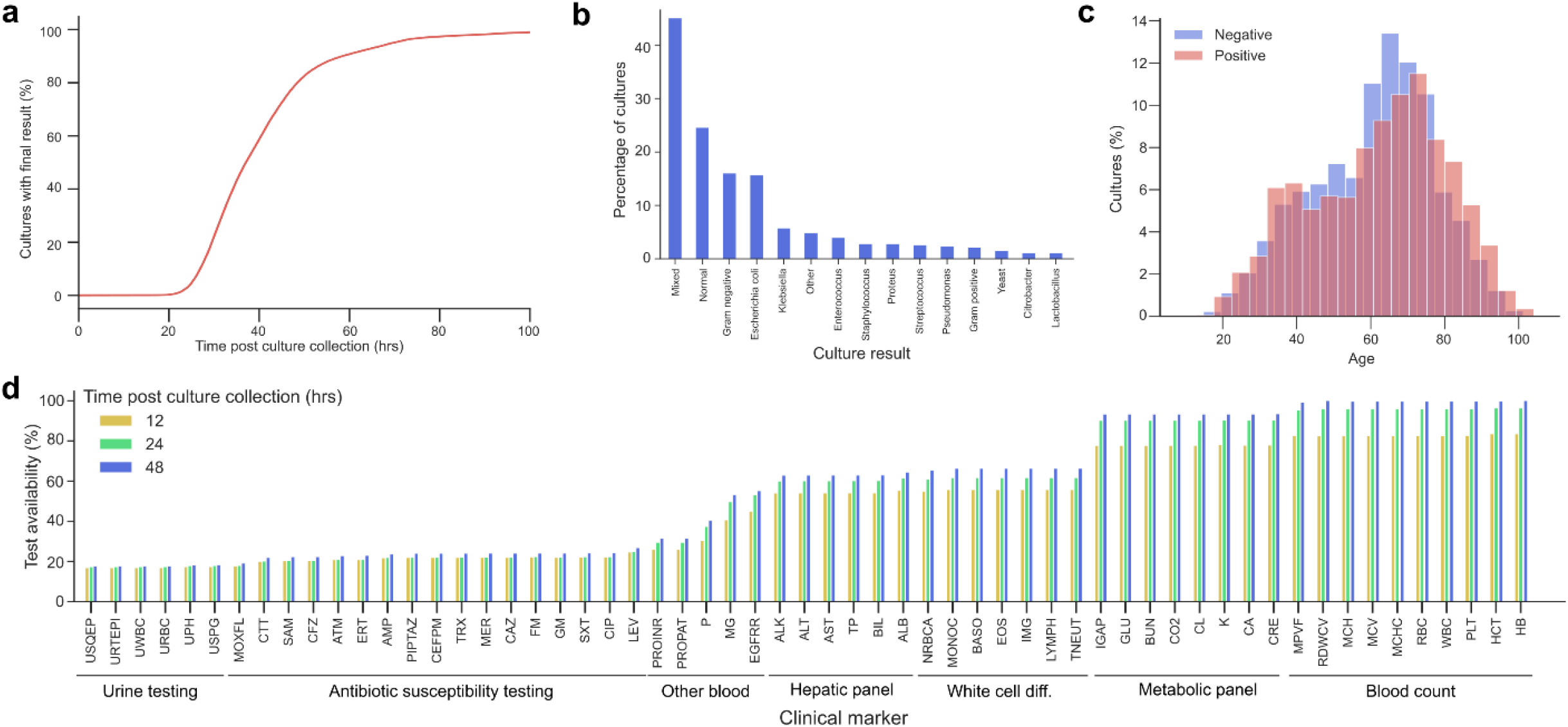
Culture result characteristics and test availability. **a**, Cumulative distribution of time taken for a culture to generate a positive finding or be finalized as negative. **b**, Distribution of positive culture findings **c**, Age distribution of positive and negative cultures. **d**, Availability of common laboratory test panels at 12-, 24-, and 48hrs post culture collection.

Of the 30,369 cultures that had associated CBC testing, >80% had this test resulted within 24hrs of the urine culture collection **(Fig. 1d)**. This suggests viability of using CBC data to predict urine culture outcomes. Availability of non-CBC testing was far more limited, with less than 20% of patients having either urine testing or antibiotic susceptibility testing results within 24hrs of the culture order.

### Prediction of urine culture results

Overall urine culture positivity could be predicted with reasonable accuracy (test set AUC: 0.76 with high overall precision (80-90%) even at high recall rates (>50%)) **(Fig. 2a)**. However, prediction of individual infectious agents exhibited mixed performance (AUCs: 0.64-0.72), generating more limited precision. Performance was quite stable over differing time periods, likely reflecting that patient test data availability did not change substantially during this window **(Fig. 2b)**. However, results were substantially improved in the subset of patients for whom urine testing was available, with AUCs of 0.8-0.85 – reflecting discriminatory capacity **(Fig. 2c)**.

**Figure 2.**
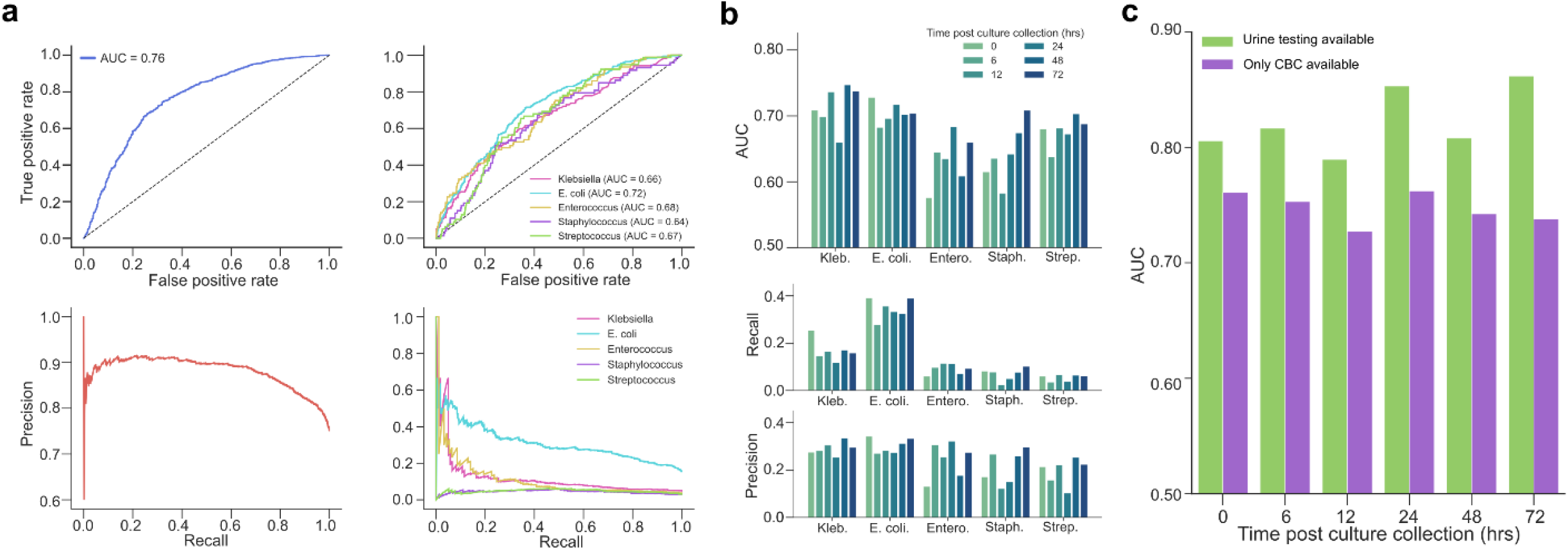
Accuracy of urine culture result. **a**, Receiver-operator characteristic (ROC) and precision-recall curves for prediction of overall culture positivity, and for specific infectious agents. **b**, Predictive accuracy over different time windows for the most common infectious agents. **c**, Area under ROC curve (AUC) with and without urine testing data available.

### Contribution of univariate features to urine culture result prediction

With age- and sex-adjusted logistic regression modelling, many features present strong and significant associations with overall culture positivity **(Fig. 3a)**. CBC, metabolic, white cell differential, hepatic, urinalysis, and antibiotic susceptibility markers all have strong outcome associations, with no one panel showing dominant associations. We see similar results when considering marker correlations with culture positivity, where correlations with individual infectious agents were far weaker **(Fig. 3b)**. Similar results were reflected in the importance of each feature to the random forest models **(Fig. 3c)**. Age, platelet count, red cell indices (HB, RBC), glucose and creatinine showed the highest overall model importance across all infectious agents. However, this result could be confounded by data availability. For example, urinalysis markers showed strong univariate correlations **(Fig. 3a-b)**, yet lacked high importance, due to lower data availability than CBC markers **(Fig. 1d)**.

**Figure 3.**
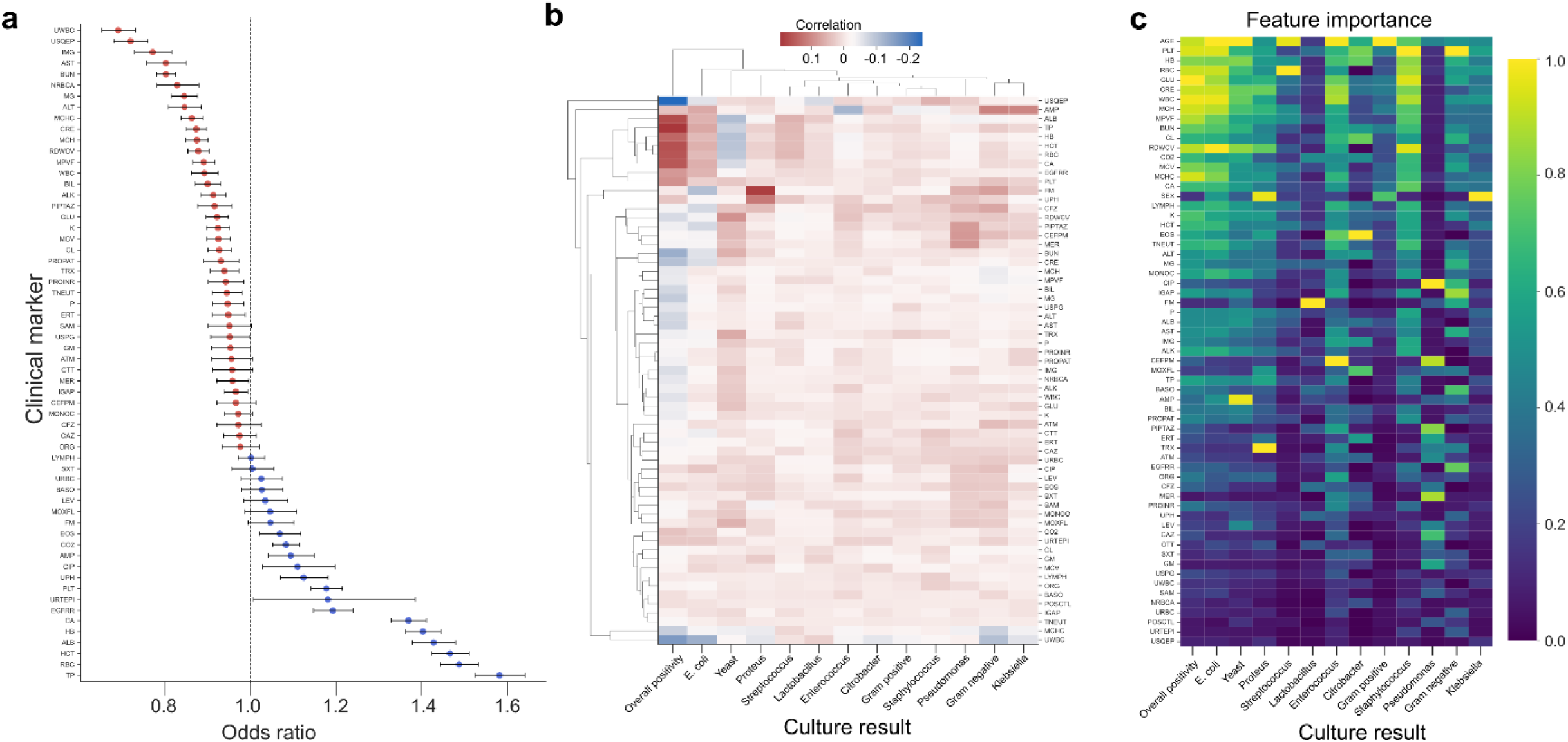
Variable associations with urine culture results. **a**, Age- and sex-adjusted odds ratios for univariate prediction of overall culture positivity. **b**, Correlation of lab tests with overall culture positivity and specific infectious agents. **c**, Machine learning model feature importance for prediction of overall culture positivity and specific infectious agents. Continuous variables in a are normalized to a 1 standard deviation increase.

## Discussion

Here we analyzed the performance of machine learning models for prediction of urine culture results using associated patient laboratory tests. While prediction of overall culture positivity was possible within a clinically valuable 24hr time window, prediction of individual infectious agents proved more challenging.

This disparity likely reflects that all major infections generate similar high-level physiologic signatures, such as increased inflammation or immune cell destruction, so inter-agent differences in these signatures may expectedly be more minor. Therefore, these differences may be obscured by the larger physiologic regulation differences of laboratory markers between patients ^15–17^.

Results in **Fig. 2c** suggest that incorporation of urinalysis results greatly improves predictive efficacy. This aligns with recent reports, where incorporation of urine dipstick test results allowed for prediction of urine culture findings with high efficacy (AUCs > 0.9)^18^. Such models may allow for targeted early detection of common infectious agents, to inform improved antibiotic usage – a high importance task in context of growing antibiotic resistance^6,7^. However, our results highlight that such performance is heavily reliant on the availability of urinalysis markers, which are not commonly collected. Across all urine cultures in our cohort, fewer than 20% had urinalysis markers within 24hrs of the culture order. This low availability makes urinalysis-based approaches infeasible for most patients, underscoring the need for models that do not rely on urinalysis, even if they achieve lower performance.

In this context, the models we present may be better suited to triage tasks, instead of directly informing treatment (e.g., antibiotic use). Given overall culture positivity prediction may be accurate enough for clinical viability, such a model could identify a subset of high-confidence negative cultures which do not require additional work-ups. This in turn could inform substantial cost savings for clinical laboratories, because of better resources allocation to the cultures more likely to be positive. Validation of our findings in additional cohorts is a clear avenue for future work.

In summary, we have developed a machine learning framework for prediction of urine culture orders using routine laboratory test markers. Our results highlight that these models can predict overall culture positivity with some efficacy, but are less performant for prediction of individual agents – particularly when urinalysis results are not available. This suggests that the primary utility of such models may be in providing rapid triage of presumptively negative cultures, to improve clinical efficiency.

## Acknowledgements

We thank the UW Department of Laboratory Medicine & Pathology Informatics team for assistance with collating UW health record data.

## Conflicts of interest

The authors have no conflicts of interest to declare.

## Data availability

Due to restrictions on sharing of protected health information, individual patient data cannot be shared.

## Author contributions

BHF conceived of and supervised the study. NH performed data collection, cleaning, and primary analysis. YC and CZ contributed to data analysis and result interpretation. All authors contributed to manuscript writing and editing.

## Notes

### Competing Interest Statement

The authors have declared no competing interest.

### Funding Statement

This study did not receive any funding

### Author Declarations

This work was performed under an ethics protocol approved by the University of Washington Institutional Review Board (STUDY00019458)

